# The post-COVID-19 population has a high prevalence of crossreactive antibodies to spikes from all *Orthocoronavirinae* genera

**DOI:** 10.1101/2023.08.01.23293522

**Authors:** Gagandeep Singh, Anass Abbad, Giulio Kleiner, Komal Srivastava, Charles Gleason, PARIS Study Group, Juan Manuel Carreño, Viviana Simon, Florian Krammer

## Abstract

The Orthocoronaviridae subfamily is large comprising four highly divergent genera. Four seasonal coronaviruses were circulating in humans prior to the coronavirus disease 2019 (COVID-19) pandemic. Infection with these viruses induced antibody responses that are relatively narrow with little cross-reactivity to spike proteins of other coronaviruses. Here, we report that infection with and vaccination against severe acute respiratory syndrome coronavirus 2 (SARS-CoV-2) induces broadly crossreactive binding antibodies to spikes from a wide range of coronaviruses including members of the sarbecovirus subgenus, betacoronaviruses including Middle Eastern respiratory syndrome coronavirus (MERS CoV), and extending to alpha-, gamma- and delta-coronavirus spikes. These data show that the coronavirus spike antibody landscape in humans has profoundly been changed and broadened as a result of the SARS-CoV-2 pandemic. While we do not understand the functionality of these crossreactive antibodies, they may lead to enhanced resistance of the population to infection with newly emerging coronaviruses with pandemic potential.

## Main text

Severe acute respiratory syndrome coronavirus 2 (SARS-CoV-2) infections and vaccinations induce binding and neutralizing antibodies to the spike protein of this new virus in humans [1, 2]. Initially, these responses led to protection from symptomatic disease as shown in a number of clinical trials. However, with the emergence of variants of concern, especially Omicron and its sub-variants, protection against symptomatic disease decreased since these new variants escaped the neutralizing antibody response induced by spike proteins from the ancestral SARS-CoV-2. However, it has been reported that binding antibody to spike protein are much better maintained against the variants as compared to neutralizing activity [3]. These binding antibodies may – in addition to T-cell immunity – contribute to the mostly maintained protection from severe disease [4].

Here, we wanted to explore how cross-reactive antibodies induced by SARS-CoV-2 infection or vaccination bind beyond the spike protein of SARS-CoV-2 and its variants. We expressed a panel of coronavirus spikes representative of all *Orthocoronavirinae* genera. We generated 21 recombinant spikes representing the five betacoronavirus (β-CoV) subgenera (sarbecoviruses, hibecoviruses, merbecoviruses, nobecoviruses and embecoviruses) as well the alphacoronavirus (α-CoV), gammacoronavirus (γ-CoV) and deltacoronavirus (δ-CoV) genera (**Figure 1A** and **Supplementary Table 1**). Using an established enzyme-linked immunosorbent assay (ELISA) [5], we tested longitudinal sera from 10 individuals who received the mRNA coronavirus disease 2019 (COVID-19) vaccine and from 10 individuals who received the vaccine after an initial SARS-CoV-2 infection **(Supplementary Table 2**). Sera were taken before vaccination, post-1^st^ dose (range 16-25 days for the vaccine-only group and 15-23 days for the infection-vaccination group) and post-2^nd^ dose (range 14-28 days for the vaccine-only group and 16-29 days for the infection-vaccination group). Binding to SARS-CoV-2 ancestral and variant spikes was induced by vaccination as expected and was detectable before vaccination in people with pre-existing immunity (**Figure 1B-D**). Comparable binding was found for the SARS-CoV-1 spike and titers were also high against three other non-SARS-CoV-2 sarbecovirus spikes tested (**Figure 1E-H**). In addition, an increase in binding was detected to a hibecovirus spike and merbecovirus spikes although at a lower level (**Figure 1I** and **L-O**). COVID-19 vaccine-associated increases were only detected for one of the two nobecovirus spikes tested (**Figure 1J** and **K**) but reactivity to embecoviruses was induced to some degree (**Figure 1P-R**) and – as expected – higher at pre-vaccination baseline since two of the spikes tested are from embecoviruses circulating in humans (OC43 and HKU1). No increase in reactivity was detected against the seasonal α-CoV spikes but pre-vaccination baseline titers to 229E spike were detectible while reactivity to NL63 spike was much lower (**Figure 1S** and **T**). Interestingly, there was also an induction of antibodies to the γ-CoV spike of HKU15 but not to the δ-CoV of HKU22 (**Figure 1U** and **V**). In general, most SARS-CoV-2 infected individuals already had titers to these spikes even before they got vaccinated. These data suggested that SARS-CoV-2 infection and vaccination can induce cross-reactive anti-spike antibodies.

**Figure 1.**
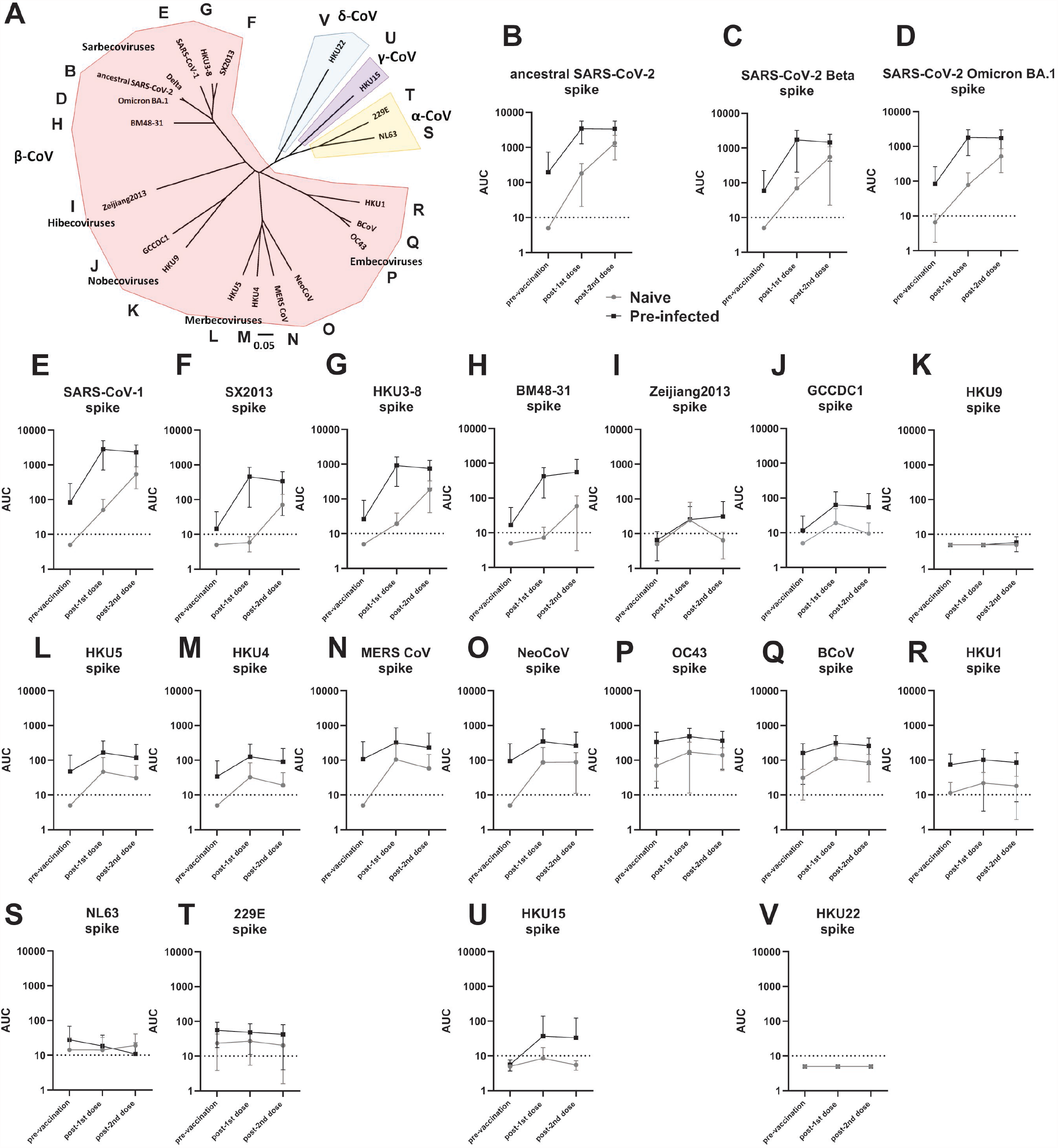
Induction of antibodies to diverse *Orthocoronavirinae* spike proteins. **A** shows a phylogenetic tree built with amino acid sequences of the spike proteins used in this study. The tree was built using Clustal Omega, visualized in FigTree and labels and highlighting was added in Microsoft Powerpoint. The scale bar indicates a 5% change in amino acid sequence. Accession numbers and full name of the different viruses strains used can be found in **Supplementary Table 1. B-D** Reactivity of longitudinal serum samples from naïve (black) or SARS-CoV-2 pre-infected (grey) individuals at a pre-vaccination time point, after the 1^st^ dose of COVID-19 mRNA vaccine and after the 2^nd^ dose of COVID-19 mRNA against the ancestral spike of SARS-CoV-2, against the Beta variant spike and against the Omicron BA.1 variant spike. **C-R** shows reactivity to diverse β–CoV spikes, **S-T** shows reactivity to α–CoV spikes, **U** shows reactivity to a δ–CoV spike and **V** shows reactivity to a γ–CoV spike. N=10 per group. Subject characteristics can be found in **Supplemental Table 2**.

We next investigated how different immune histories influence pan-coronavirus seroreactivity. We tested sera from different exposure groups including pre-pandemic samples (n=15, collected between 2018-2019), samples from convalescent individuals (n=16, collected at 23-87 days post infection), individuals who got two doses of COVID-19 mRNA vaccines (n=20, range of 14-36 days post-2^nd^ dose), individuals who got three doses of COVID-19 mRNA vaccines (n=20, range of 14-33 days post-3^rd^ dose), individuals who were infected and then got two doses of COVID-19 mRNA vaccines (n=20, range of 15-39 days post-2^nd^ dose) and individuals who got infected and three doses of COVID-19 mRNA vaccines (n=9, range of 14-30 days post-3^rd^ dose). More information regarding the specific samples can be found in **Supplementary Table 3**. Analysis of pre-pandemic sera showed reactivity to the β-CoV spikes from two human seasonal coronaviruses, OC43 and HKU1. Interestingly, there was also strong reactivity, comparable to OC43, to the spike of the bovine coronavirus (BCoV) with sera from all study participants having reactivity above the limit of detection (LoD) (**Figure 2A, Extended Figure 1A**). Reactivity against spikes from seasonal α-CoVs, NL63 and 229E was also found. Similar to data shown in **Figure 1S** and **T**, reactivity to NL63 was much lower than for 229E. For sera from SARS-CoV-2 convalescent individuals, we observed strong reactivity to SARS-CoV-2 spikes with elevated reactivity to spikes of many β-CoV with the exception of the nobecovirus spikes from GCCDC1 and HKU9 (**Figure 2B**). Sera from several study particpants (44% and 56% respectively) even had detectible reactivity to the γ-CoV spike of HKU15 and δ-CoV spike of HKU22 (**Extended Figure 1B**). By receiving two or three doses of COVID-19 mRNA vaccines the reactivity pattern changed slightly. With two doses of the vaccine, higher reactivity to β-CoV spikes could be observed (**Figure 2C**). With three doses of the mRNA vaccine, high and relatively uniform reactivity to sarbecoviruses can be measured (**Figure 2D**). In addition, the percentage of individuals who had detectible reactivity in the 2x and 3x mRNA vaccinated groups was high across all tested spikes (**Extended Figure 1C** and **D**). Sera from people who had received three vaccine doses cross-reacted with all spikes except the spikes of the the nobecoviruses GCCDC1 and HKU9 and the γ-CoV spike of HKU15 and to the δ-CoV of HKU22 where about approximately 20-55% of individuals had reactivity (**Extended Figure 1D**). Similarly strong reactivity across the board was also detected in individuals who had been infected and then vaccinated 2x or 3x with COVID-19 mRNA vaccines (**Figures 2E** and **F, Extended Figure 1E** and **F**).

**Figure 2.**
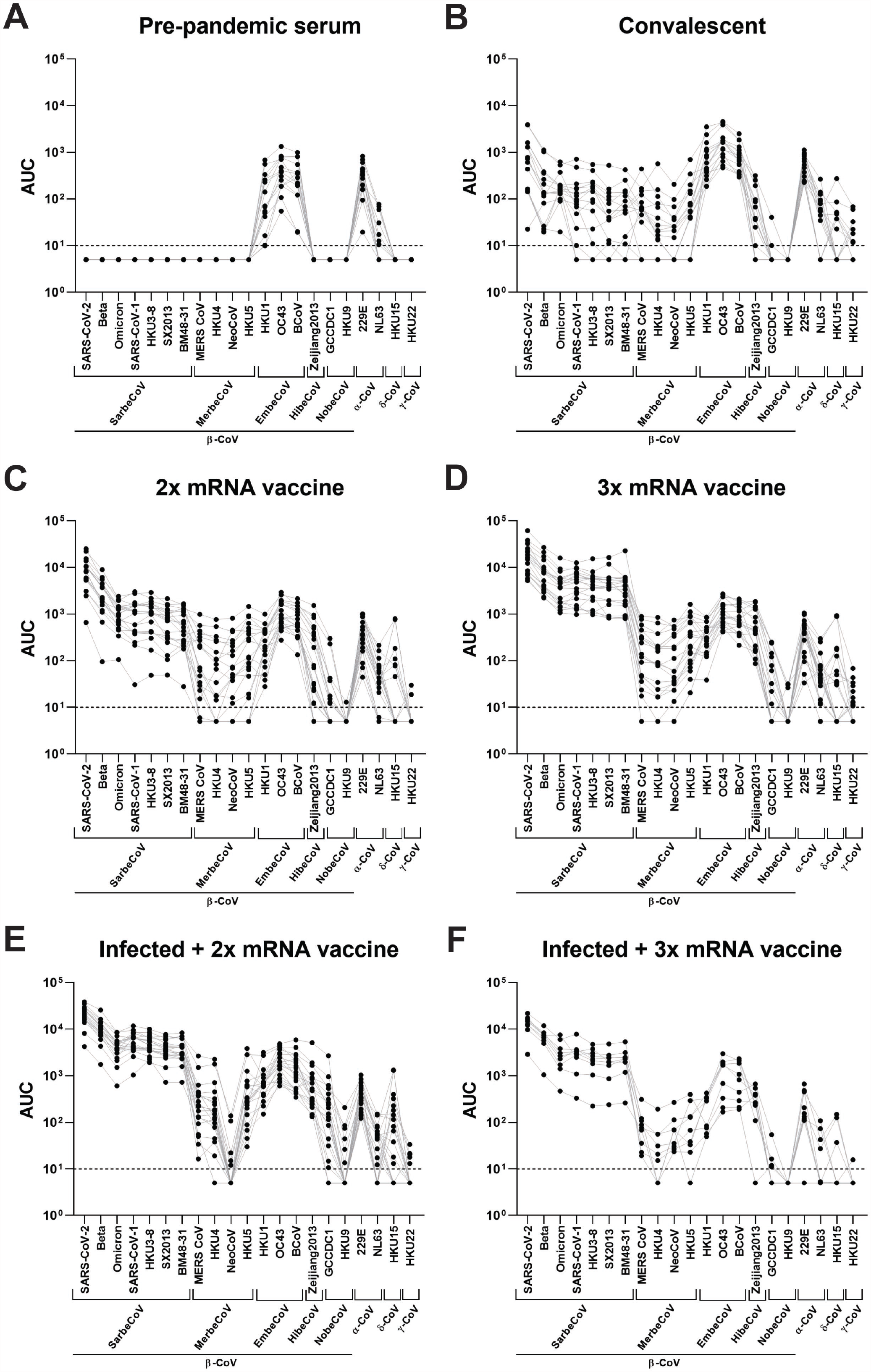
Cross-sectional reactivity of human sera from different exposure groups to diverse *Orthocoronavirinae* spike proteins. **A** shows serum reactivity of pre-pandemic serum sample to different spike proteins (n=15). **B** shows serum reactivity of SARS-CoV-2 convalescent individuals to different spike proteins (n=16). **C** shows serum reactivity of individuals vaccinated 2x with COVID-19 mRNA vaccines, **D** shows serum reactivity of individuals who got three doses of vaccine (n=20 for each group) to different spike proteins. **E** and **F** show serum reactivity of individuals infected with SARS-CoV-2 and then vaccinated twice (n=20 except for SX2013 where n=19) or three times (n=9) to different spike proteins. Subject characteristics can be found in **Supplemental Table 3**.

Several of our observations from these experiments are interesting. Even in pre-COVID-19 sera, the prevalence and titers against the spike of β-CoV BCoV are almost as high as against the spike of seasonal β-CoV OC43. This indicates an antigenic relatedness of the BCoV and OC43 spikes. It has been hypothesized that OC43 shares a common ancestor with BCoV and may have split off around 1890 [6]. This split would coincide with the ‘Russian flu’ pandemic of 1889/1890 and it has been hypothesized that this pandemic was, indeed, caused by an OC43 ancestor which jumped from cattle into humans [7]. The crossreactivity found may perhaps further support this hypothesis and also explain why zoonotic infections in humans with BCoV are rare even though the virus can infect children [8]. In addition, the titers as well as the percentage of positive individuals for the seasonal α-CoV 229E are much higher than for the seasonal α-CoV NL63. These differences may point perhaps to an inherent difference in immunogenicity, antibody durability or represent an artifact of the protein used in the ELISA. Furthermore, from our longitudinal sample set, we noted that COVID-19 mRNA vaccination induces antibodies to diverse spike proteins including many β-CoVs spikes as well as δ-CoV spikes. As previously reported, backboosting to OC43 and HKU (and BCoV) was also observed [9, 10], but little to no induction of antibodies was seen against α-CoV spikes, the tested γ-CoV spike or the spikes from the nobecoviruses subgenus of the β-CoV. At a cross-sectional level we observed that sera from individuals infected with SARS-CoV-2 already show relatively broad reactivity across β-CoVs spikes with about half of the individuals having also some reactivity to δ-CoV and γ-CoV spikes. COVID-19 mRNA vaccination induced similar crossreactivity. However, especially after three vaccine doses, there seems to be a bias towards sarbecoviruses with a clear drop towards other non-seasonal β-CoV spikes. It is important to note that after mRNA vaccination with the original vaccine containing the ancestral SARS-CoV-2 spike, titers against diverse sarbecovirus spikes are as high as against the Omicron spike. This may suggest, that the protection from severe disease that we observe against Omicron sublineages perhaps also applies to other, diverse sabecoviruses (e.g., SARS-CoV-1, HKU3-8, SX2013, BM48-31) which may (re)-emerge in the future as human pathogens. While these cross-reactive antibodies may not neutralize more distantly related viruses, they may still afford protection via Fc-mediated effector functions.

Another interesting aspect is the question of which epitopes these crossreactive antibodies target. Crossreactivity within the sarbecoviruses likely targets the receptor binding domain RBD but also different epitopes on the S2 subunit. Cross-reactivity to other β-CoV spikes is likely mediated mostly by S2 targeting antibodies since the receptor binding domain (RBD) is probably too divergent. Such antibodies have been isolated with some of them having neutralizing activity although at low potency [9, 11-13]. Finally, crossreactivity to α-CoV, δ-CoVs and γ-CoVs is likely due to more rare antibodies that target the fusion peptide in the S2 domain. These antibodies have also been isolated and they exert neutralizing activity to some extend [14, 15]. However, it would be helpful to isolate more mAbs with different reactivity profiles to understand their characteristics better and determine if they indeed can contribute to protection *in vivo*. Of note, monoclonal antibody therapeutics and prophylactics have been very efficient treatment options for COVID-19, but antigenic changes of SARS-CoV-2 have rendered them irrelevant. Perhaps antibodies that target more conserved epitopes, even if they have lower neutralizing potency, could be a more sustainable solution.

In summary, we found a high prevalence of antibodies that crossreact to spike proteins from all four *Orthocoronavirinae* genera. It is entirely possible that the global population, which is hyperimmunized to SARS-CoV-2 through infection and vaccination, has now build more resistance to the many members coronavirus family in general.

## Data Availability

All data produced in the present study are available upon reasonable request to the authors.

## Acknowledgements

We thank all the participants of our longitudinal PARIS study for their generous and continued support of research. We would like to thank Dr. D. Noah Sather from the University of Washington for providing the Beta and BA.1 spike protein used in this study and Drs. Kizzmekia Corbett and Barney Graham for providing 229E, NL63, HKU1 and OC43 constructs. This effort was supported by the Serological Sciences Network (SeroNet) in part with Federal funds from the National Cancer Institute, National Institutes of Health, under Contract No. 75N91019D00024, Task Order No. 75N91021F00001. The content of this publication does not necessarily reflect the views or policies of the Department of Health and Human Services, nor does mention of trade names, commercial products or organizations imply endorsement by the U.S. Government. This work was also partially funded by the Centers of Excellence for Influenza Research and Surveillance (CEIRS, contract # HHSN272201400008C), the Centers of Excellence for Influenza Research and Response (CEIRR, contract # 75N93021C00014), by the Collaborative Influenza Vaccine Innovation Centers (CIVICs contract # 75N93019C00051) and by institutional funds.

## Online Methods

### Recombinant proteins expression

Mammalian expression vectors encoding the ectodomain of spike proteins from SARS-CoV-1 (GenBank: 875 AAP13441.1), SARS-CoV-2 (GenBank: MN908947.3), HKU3-8 (GenBank: ADE34766.1), SX2013 (GenBank: AIA62300.1), BM48-31(GenBank: YP_003858584.1), SARS-CoV-2 Omicron (GenBank: UFT26501.1), MERS-CoV (GenBank: AXP07355.1), HKU4 (GenBank: YP_001039953.1), HKU5 (GenBank: YP_001039962.1), HKU9 (YP_001039971.1), GCCDC1 (GenBank: QKF94914.1), Zhejiang2013, bat Hp (GenBank: YP_009072440.1) HKU15 (GenBank: YP_009513021.1), HKU22 (GenBank: AHB63508.1), BCoV, bovine coronavirus (GenBank: AAA66399.1), Neo CoV, Coronavirus Neoromicia (Genbank: AGY29650.2), 229E (Genbank: NP_073551.1), NL63 (GenBank AFV53148.1), OC43 (Genbank: KF963240.1) and HKU1 (Genbank: AGW27881.1) (see **Supplementary Table 1** for more information on virus isolates, host and receptor) with a C-terminal thrombin cleavage site, T4 foldon trimerization domain and hexahistidine tag were constructed as described earlier [5, 16]. The constructs also contain the “2P” stabilizing mutations [17] and known or putative S1-S2 cleavage sites were removed. Proteins were purified from transiently-transfected Expi293F cells with each respective plasmid. Cell-free supernatant was harvested after 3 days post transfection and his-tagged proteins were purified by gravity chromatography using Ni^2+^-nitriloacetic acid (NTA) agarose (Qiagen). Proteins were eluted and buffer was exchanged using Amicon centrifugal units (EMD Millipore) and all recombinant proteins were finally re-suspended in phosphate buffered saline (PBS) as described [16]. Proteins were run on sodium dodecyl sulfate polyacrylamide gel electrophoresis (SDS-PAGE) gels under reducing conditions for quality control and stored at -80°C until use.

### ELISA

Antibody titers in in serum samples were assessed using a research grade ELISA [5, 16] using recombinant versions of full-length spike (S) proteins of different coronaviruses. Briefly, Immulon 4 HBX 96-well microtiter plates (Thermo Fisher Scientific) were coated overnight at 4 °C with 50 μl per well of a 2 μg/ml solution of each respective recombinant protein resuspended in PBS (Gibco; cat. no. 10010-031). The next morning, plates were washed 3 times with PBS supplemented with 0.1% Tween-20 using an automatic plate washer (BioTek 405TS microplate washer) and blocked with 200 μl per well of PBST containing 3% milk powder (AmericanBio) for 1 h at room temperature (RT). Blocking solution was removed, and initial dilutions (1:100) of heat-inactivated serum (in PBS-T 1% milk powder) were added to the plates, followed by 2-fold serial dilutions and 2h incubation at RT. Next, plates were washed three times with PBS-T and 50 μl per well of the pre-diluted secondary anti-human IgG (Fab-specific) horseradish peroxidase (HRP) antibody (produced in goat; Sigma-Aldrich, cat. no. A0293, RRID: AB_257875) diluted 1:9,000 in PBS-T containing 1% milk powder were added for 1 h. Plates were again washed three times with 0.1% PBST, the substrate o-phenylenediamine dihydrochloride (OPD) (SIGMAFAST) was added (100 μl per well) for 10 min, followed by an addition of 3M hydrochloric acid (50 μl per well; Thermo Fisher Scientific) to stop the reaction. Optical density (OD) was measured at a wavelength of 490 nm using a plate reader (BioTek, Synergy H1 microplate reader). The area under the curve (AUC) values were calculated and plotted using Prism 9 software (GraphPad).

### Human serum samples

Human serum samples were obtained from study participants in the longitudinal observational Protection Associated with Rapid Immunity to SARS-CoV-2 (PARIS) study [18]. This cohort follows health care workers longitudinally since April 2020. The study was reviewed and approved by the Icahn school of Medicine at Mount Sinai Institutional Review Board (IRB-20-03374). All participants provided informed consent and HIPAA Authorization prior to sample and data collection. All participants provided permission for sample banking and sharing. The participants did not receive compensation. All biospecimen were coded and stored at −80 °C.

We used longitudinal serum samples collected from 20 adult study participants. 10/20 of the study participants (50%) were infected with SARS-CoV-2 prior to the first vaccine dose and were seropositive prior to vaccination (pre-infected group). 10/20 study participants (50%) had no previous SARS-CoV-2 infection history and were seronegative for SARS-CoV-2 antibodies prior to vaccination (naive group). Participants received two doses of either the Moderna mRNA-1273 vaccine or the Pfizer-BioNTech BNT162b2 vaccine. Demographics of seropositive and seronegative study participants and sample collection time points from each individual are summarized in **Supplementary Table 2**.

For the antigenic landscape characterization against various coronaviruses, we selected 85 serum samples from 54 participants. 20 out of 54 participants were seronegative prior to vaccination while 34/54 had COVID-19 prior to vaccination. All participants with pre-vaccination immunity were infected in 2020 when only ancestral SARS-CoV-2 strains circulated in the New York metropolitan area. Convalescent samples (n = 15) were obtained within three months of SARS-CoV-2 infection (average: 58 days, range: 23–87 days) whereas the post-vaccination samples were collected on average 23 days (range: 14–39 days) after the second dose [n = 40; where n = 20 Pfizer 2×(10 individuals with prior infection and 10 individuals with no infection) and n = 20 Moderna 2× (10 individuals with prior infection and 10 individuals with no infection)] or 19 days (range: 14–33 days) after the third booster [n = 30; 20 Pfizer 3× (10 individuals with prior infection and 10 individuals with no infection) and 10 Moderna 3×) vaccine dose. Pre-pandemic human serum samples were collected before the COVID-19 pandemic. Demographics of participants and sample collection time points from each individual are summarized in **Supplementary Table 3**.

## Supplementary Tables

**Supplementary Table 1:** Information on the viral isolates used for recombinant spike protein generation.

| Virus | Genus (subgenus) | Accession # | Host Species | Receptor | Reference |
| --- | --- | --- | --- | --- | --- |
| 229E | α-CoV | NP_073551.1 | *Homo sapiens* | Human aminopeptidase N (APN) | [19] |
| NL63 | α-CoV | AFV53148.1 | *Homo sapiens* | Angiotensin converting enzyme 2 (ACE2) | [20] |
| SARS CoV-2 | β-CoV (sarbecovirus) | MN908947.3 | *Homo sapiens* | ACE2 | [21] |
| SARS-CoV-1 | β-CoV (sarbecovirus) | AAP13441.1 | Several species | ACE2 | [22] |
| HKU3-8 | β-CoV (sarbecovirus) | ADE34766.1 | *Rhinolophus* species | | [23] |
| BM48-31 | β-CoV<br>(sarbecovirus) | YP_003858584.1 | *Rhinolophus blasii* | | [23] |
| SX2013 | β-CoV<br>(sarbecovirus) | AIA62300.1 | *Rhinolophus ferrumequinum* | | [23] |
| MERS CoV | β-CoV<br>(merbecovirus) | AXP07355.1 | Camelus dromedariu | Dipeptidyl peptidase 4 (DPP4) | [24] |
| HKU4 | β-CoV<br>(merbecovirus) | YP_001039953.1 | *Tylonycteris* species | DDP4 | [25, 26] |
| HKU5 | β-CoV<br>(merbecovirus) | YP_001039962.1 | *Pipistrellus* species | | [25, 27] |
| NeoCoV | β-CoV<br>(merbecovirus) | AGY29650.2 | Neoromicia capensis | ACE2 | [28, 29] |
| HKU9 | β-CoV<br>(nobecovirus) | YP_001039971.1 | *Rousettus leschenaulti* | | [27] |
| GCCDC 1 | β-CoV<br>(nobecovirus) | QKF94914.1 | *Rousettus leschenaulti* | | [30] |
| Zhejiang2013 (Bat Hp-betacoronavirus/Zhejiang2013) | β-CoV<br>(hibecovirus) | YP_009072440.1 | *Hipposideros pratti* | | [31] |
| BCoV | β-CoV<br>(embecovirus) | AAA66399.1 | Bovine species | N-acetyl-9-O-acetylneuraminic acid (Neu5,9Ac2). | [32] |
| OC43 | β-CoV<br>(embecovirus) | KF963240.1 | *Homo sapiens sapiens* | N-acetyl-9-O-acetylneuraminic acid (Neu5,9Ac2). | [33] |
| HKU1 | β-CoV<br>(embecovirus) | AGW27881.1 | *Homo sapiens sapiens* | N-acetyl-9-O-acetylneuraminic acid (Neu5,9Ac2). | [33] |
| HKU15 | δ-CoV | YP_009513021.1 | *Sus scrofa* | porcine aminopeptidase N (pAPN) | [34] |
| HKU22 | γ-CoV | AHB63508.1 | *Tursiops*<br>species |  | [35] |

**Supplementary Table 2:** Characteristics of individuals shown in Figure 1.

| **Participant ID** | **Age Bracket** | **Sex** | **Time points included in this study** | **SARS CoV-2 infection prior to vaccination** | **Vaccine type** |
| --- | --- | --- | --- | --- | --- |
| Pluto-001 | 45-49 | Male | Pre vaccine, Post-vax, Post-Boost | Yes | Pfizer |
| Pluto-002 | 20-24 | Female | Pre vaccine, Post-vax, Post-Boost | Yes | Moderna |
| Pluto-003 | 55-59 | Female | Pre vaccine, Post-vax, Post-Boost | No | Moderna |
| Pluto-004 | 50-54 | Female | Pre vaccine, Post-vax, Post-Boost | Yes | Pfizer |
| Pluto-005 | 40-44 | Female | Pre vaccine, Post-vax, Post-Boost | No | Moderna |
| Pluto-006 | 35-39 | Female | Pre vaccine, Post-vax, Post-Boost | No | Pfizer |
| Pluto-007 | 65-69 | Male | Pre vaccine, Post-vax, Post-Boost | No | Moderna |
| Pluto-008 | 50-54 | Female | Pre vaccine, Post-vax, Post-Boost | Yes | Pfizer |
| Pluto-009 | 25-29 | Female | Pre vaccine, Post-vax, Post-Boost | Yes | Pfizer |
| Pluto-0010 | 25-29 | Female | Pre vaccine, Post-vax, Post-Boost | No | Moderna |
| Pluto-0011 | 60-64 | Male | Pre vaccine, Post-vax, Post-Boost | Yes | Pfizer |
| Pluto-0012 | 25-29 | Male | Pre vaccine, Post-vax, Post-Boost | No | Moderna |
| Pluto-0013 | 25-29 | Male | Pre vaccine, Post-vax, Post-Boost | Yes | Pfizer |
| Pluto-0014 | 35-39 | Female | Pre vaccine, Post-vax, Post-Boost | Yes | Moderna |
| Pluto-0015 | 20-24 | Female | Pre vaccine, Post-vax, Post-Boost | No | Pfizer |
| Pluto-0016 | 25-29 | Female | Pre vaccine, Post-vax, Post-Boost | Yes | Pfizer |
| Pluto-0017 | 40-44 | Female | Pre vaccine, Post-vax, Post-Boost | No | Moderna |
| Pluto-0018 | 40-44 | Female | Pre vaccine, Post-vax, Post-Boost | No | Pfizer |
| Pluto-0019 | 30-34 | Female | Pre vaccine, Post-vax, Post-Boost | No | Pfizer |
| Pluto-0020 | 35-39 | Male | Pre vaccine, Post-vax, Post-Boost | Yes | Moderna |

**Supplementary Table 3:** Characteristics of individuals shown in Figure 2 and S Figure 1.

| **Participant ID** | **Age Bracket** | **Sex** | **Time points included in this study** | **SARS CoV-2 infection prior to vaccination** | **Vaccine Type** |
| --- | --- | --- | --- | --- | --- |
| OMI-001 | 30-39 | Female | Post-Vax, Post-Boost | No | Moderna |
| OMI-002 | 18-29 | Female | Post-Vax, Post-Boost | No | Moderna |
| OMI-003 | 40-49 | Male | Post-Vax, Post-Boost | No | Moderna |
| OMI-004 | 40-49 | Female | Post-Vax, Post-Boost | No | Moderna |
| OMI-005 | 40-49 | Female | Post-Vax, Post-Boost | No | Moderna |
| OMI-006 | 40-49 | Male | Post-Vax, Post-Boost | No | Moderna |
| OMI-007 | 50-59 | Female | Post-Vax, Post-Boost | No | Moderna |
| OMI-008 | 60-69 | Female | Post-Vax, Post-Boost | No | Moderna |
| OMI-009 | 30-39 | Female | Post-Vax, Post-Boost | No | Moderna |
| OMI-010 | 18-29 | Female | Post-Vax, Post-Boost | No | Moderna |
| OMI-011 | 40-49 | Female | Post-Vax, Post-Boost | No | Pfizer |
| OMI-012 | 18-29 | Female | Post-Vax, Post-Boost | No | Pfizer |
| OMI-013 | 40-49 | Female | Post-Vax, Post-Boost | No | Pfizer |
| OMI-014 | 50-59 | Female | Post-Vax, Post-Boost | No | Pfizer |
| OMI-015 | 18-29 | Female | Post-Vax, Post-Boost | No | Pfizer |
| OMI-016 | 70-79 | Male | Post-Vax, Post-Boost | No | Pfizer |
| OMI-017 | 30-39 | Female | Post-Vax, Post-Boost | No | Pfizer |
| OMI-018 | 30-39 | Female | Post-Vax, Post-Boost | No | Pfizer |
| OMI-019 | 40-29 | Male | Post-Vax, Post-Boost | No | Pfizer |
| OMI-020 | 40-49 | Female | Post-Vax, Post-Boost | No | Pfizer |
| OMI-021 | 40-49 | Female | Post-Infection, Post-Vax | Yes | Pfizer |
| OMI-022 | 30-39 | Female | Post-Infection, Post-Vax | Yes | Pfizer |
| OMI-023 | 30-39 | Female | Post-Infection, Post-Vax | Yes | Pfizer |
| OMI-024 | 30-39 | Male | Post-Infection, Post-Vax | Yes | Pfizer |
| OMI-028 | 18-29 | Female | Post-Infection, Post-Vax | Yes | Moderna |
| OMI-029 | 30-39 | Female | Post-Infection, Post-Vax | Yes | Pfizer |
| OMI-032 | 50-59 | Male | Post-Infection, Post-Vax | Yes | Pfizer |
| OMI-027 | 40-49 | Female | Post-Infection, Post-Vax | Yes | Pfizer |
| OMI-033 | 30-39 | Female | Post-Infection, Post-Vax | Yes | Pfizer |
| OMI-034 | 40-49 | Male | Post-Infection, Post-Vax | Yes | Pfizer |
| OMI-035 | 30-39 | Female | Post-Infection, Post-Vax | Yes | Pfizer |
| OMI-025 | 50-59 | Male | Post-Infection | Yes | No vax |
| OMI-026 | 30-39 | Male | Post-Infection | Yes | No vax |
| OMI-030 | 40-49 | Female | Post-Infection | Yes | No vax |
| OMI-031 | 30-39 | Female | Post-Infection | Yes | No vax |
| OMI-036 | 50-59 | Male | Post-Vax | Yes | Moderna |
| OMI-037 | 30-39 | Male | Post-Vax | Yes | Moderna |
| OMI-038 | 18-29 | Male | Post-Vax | Yes | Moderna |
| OMI-039 | 30-39 | Female | Post-Vax | Yes | Moderna |
| OMI-040 | 60-69 | Female | Post-Vax | Yes | Moderna |
| OMI-041 | 40-49 | Female | Post-Vax | Yes | Moderna |
| OMI-042 | 40-49 | Female | Post-Vax | Yes | Moderna |
| OMI-043 | 60-69 | Male | Post-Vax | Yes | Moderna |
| OMI-044 | 40-49 | Male | Post-Vax | Yes | Moderna |
| OMI-045 | 30-39 | Male | Post-Boost | Yes | Pfizer |
| OMI-046 | 30-39 | Female | Post-Boost | Yes | Pfizer |
| OMI-047 | 50-59 | Female | Post-Boost | Yes | Pfizer |
| OMI-048 | 60-69 | Male | Post-Boost | Yes | Pfizer |
| OMI-049 | 60-69 | Male | Post-Boost | Yes | Pfizer |
| OMI-050 | 30-39 | Female | Post-Boost | Yes | Pfizer |
| OMI-051 | 50-59 | Female | Post-Boost | Yes | Pfizer |
| OMI-052 | 50-59 | Female | Post-Boost | Yes | Pfizer |
| OMI-053 | 18-29 | Female | Post-Boost | Yes | Pfizer |
| OMI-054 | 30-39 | Female | Post-Boost | Yes | Pfizer |

**Extended Figure 1:**
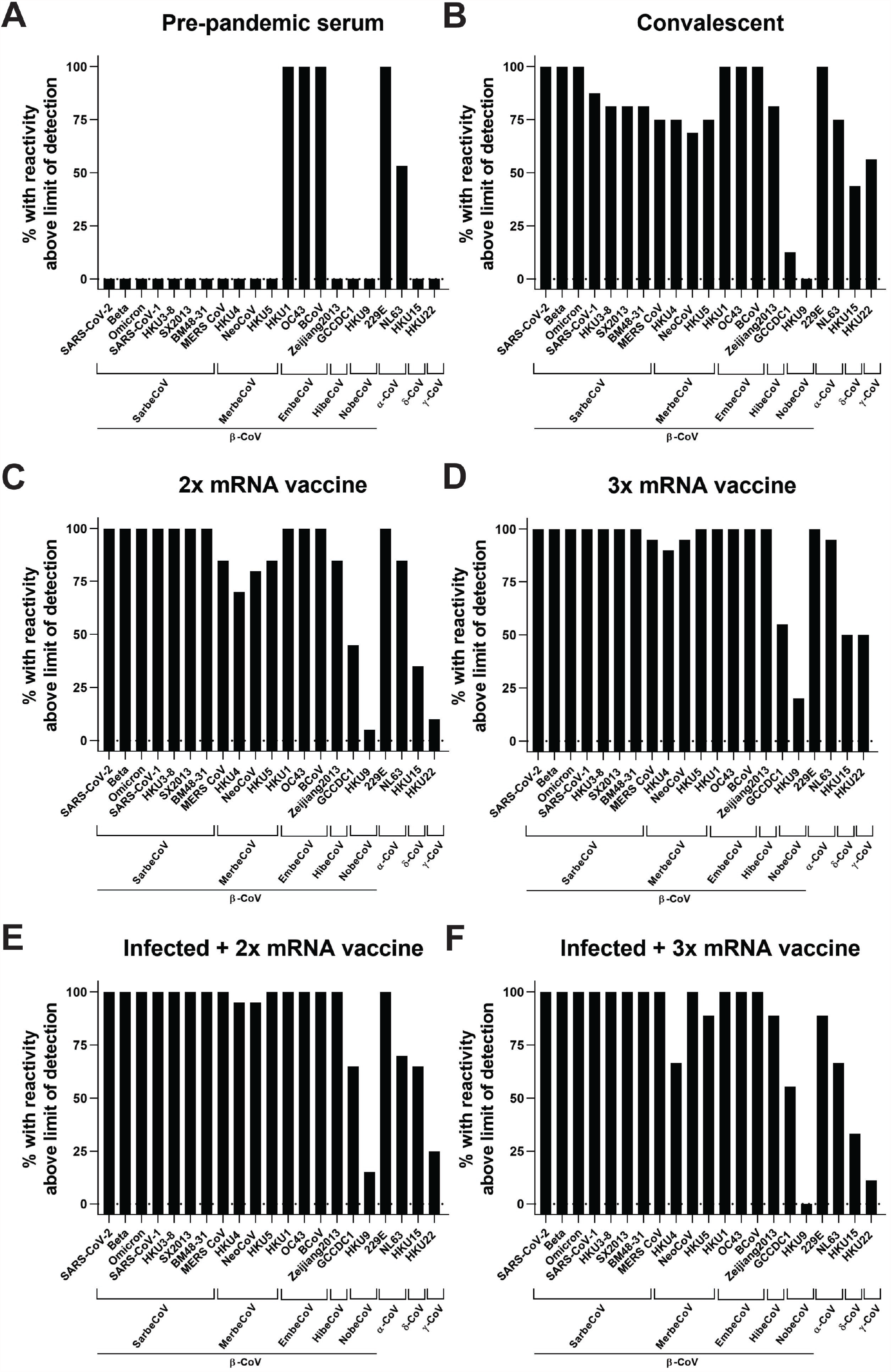
Percent of individuals with reactivity above the limit of detection (LoD) to different spike proteins for each group shown in Figure 1. **A** shows percentage of pre-pandemic serum samples reactive to different spike proteins (n=15). **B** shows percentage of samples from SARS-CoV-2 convalescent individuals reactive to different spike proteins (n=16). **C** shows percentage of individuals vaccinated 2x with COVID-19 mRNA vaccines reactive to different spikes, **D** shows the same for individuals who got three doses of vaccine (n=20 for each group). **E** and **F** show percentage of individuals infected with SARS-CoV-2 and then vaccinated twice (n=20 except for SX2013 where n=19) or three times (n=9) who had reactivity to different spike proteins. Subject characteristics can be found in **Supplemental Table 3**.

## References

[1] Krammer F, Srivastava K, Alshammary H, Amoako AA, Awawda MH, Beach KF, et al. Antibody Responses in Seropositive Persons after a Single Dose of SARS-CoV-2 mRNA Vaccine. N Engl J Med. 2021.

[2] Wajnberg A, Amanat F, Firpo A, Altman DR, Bailey MJ, Mansour M, et al. Robust neutralizing antibodies to SARS-CoV-2 infection persist for months. Science. 2020;370:1227–30.

[3] Carreño JM, Alshammary H, Tcheou J, Singh G, Raskin A, Kawabata H, et al. Activity of convalescent and vaccine serum against SARS-CoV-2 Omicron. Nature. 2021.

[4] Feikin DR, Abu-Raddad LJ, Andrews N, Davies MA, Higdon MM, Orenstein WA, et al. Assessing vaccine effectiveness against severe COVID-19 disease caused by omicron variant. Report from a meeting of the World Health Organization. Vaccine. 2022;40:3516–27.

[5] Amanat F, Stadlbauer D, Strohmeier S, Nguyen THO, Chromikova V, McMahon M, et al. A serological assay to detect SARS-CoV-2 seroconversion in humans. Nat Med. 2020.

[6] Vijgen L, Keyaerts E, Moës E, Thoelen I, Wollants E, Lemey P, et al. Complete genomic sequence of human coronavirus OC43: molecular clock analysis suggests a relatively recent zoonotic coronavirus transmission event. J Virol. 2005;79:1595–604.

[7] Brüssow H, Brüssow L. Clinical evidence that the pandemic from 1889 to 1891 commonly called the Russian flu might have been an earlier coronavirus pandemic. Microb Biotechnol. 2021;14:1860–70.

[8] Zhang XM, Herbst W, Kousoulas KG, Storz J. Biological and genetic characterization of a hemagglutinating coronavirus isolated from a diarrhoeic child. J Med Virol. 1994;44:152–61.

[9] Amanat F, Thapa M, Lei T, Ahmed SMS, Adelsberg DC, Carreño JM, et al. SARS-CoV-2 mRNA vaccination induces functionally diverse antibodies to NTD, RBD, and S2. Cell. 2021;184:3936–48.e10.

[10] Aydillo T, Rombauts A, Stadlbauer D, Aslam S, Abelenda-Alonso G, Escalera A, et al. Immunological imprinting of the antibody response in COVID-19 patients. Nat Commun. 2021;12:3781.

[11] Zhou P, Song G, Liu H, Yuan M, He WT, Beutler N, et al. Broadly neutralizing anti-S2 antibodies protect against all three human betacoronaviruses that cause deadly disease. Immunity. 2023;56:669–86.e7.

[12] Pinto D, Sauer MM, Czudnochowski N, Low JS, Tortorici MA, Housley MP, et al. Broad betacoronavirus neutralization by a stem helix-specific human antibody. Science. 2021;373:1109–16.

[13] Song G, He WT, Callaghan S, Anzanello F, Huang D, Ricketts J, et al. Cross-reactive serum and memory B-cell responses to spike protein in SARS-CoV-2 and endemic coronavirus infection. Nat Commun. 2021;12:2938.

[14] Dacon C, Tucker C, Peng L, Lee CD, Lin TH, Yuan M, et al. Broadly neutralizing antibodies target the coronavirus fusion peptide. Science. 2022;377:728–35.

[15] Low JS, Jerak J, Tortorici MA, McCallum M, Pinto D, Cassotta A, et al. ACE2-binding exposes the SARS-CoV-2 fusion peptide to broadly neutralizing coronavirus antibodies. Science. 2022;377:735–42.

[16] Stadlbauer D, Amanat F, Chromikova V, Jiang K, Strohmeier S, Arunkumar GA, et al. SARS-CoV-2 Seroconversion in Humans: A Detailed Protocol for a Serological Assay, Antigen Production, and Test Setup. Curr Protoc Microbiol. 2020;57:e100.

[17] Pallesen J, Wang N, Corbett KS, Wrapp D, Kirchdoerfer RN, Turner HL, et al. Immunogenicity and structures of a rationally designed prefusion MERS-CoV spike antigen. Proc Natl Acad Sci U S A. 2017;114:E7348–E57.

[18] Simon V, Kota V, Bloomquist RF, Hanley HB, Forgacs D, Pahwa S, et al. PARIS and SPARTA: Finding the Achilles’ Heel of SARS-CoV-2. mSphere. 2022;7:e0017922.

[19] Yeager CL, Ashmun RA, Williams RK, Cardellichio CB, Shapiro LH, Look AT, et al. Human aminopeptidase N is a receptor for human coronavirus 229E. Nature. 1992;357:420–2.

[20] Hofmann H, Pyrc K, van der Hoek L, Geier M, Berkhout B, Pöhlmann S. Human coronavirus NL63 employs the severe acute respiratory syndrome coronavirus receptor for cellular entry. Proc Natl Acad Sci U S A. 2005;102:7988–93.

[21] Letko M, Marzi A, Munster V. Functional assessment of cell entry and receptor usage for SARS-CoV-2 and other lineage B betacoronaviruses. Nat Microbiol. 2020;5:562–9.

[22] Li W, Moore MJ, Vasilieva N, Sui J, Wong SK, Berne MA, et al. Angiotensin-converting enzyme 2 is a functional receptor for the SARS coronavirus. Nature. 2003;426:450–4.

[23] Wells HL, Letko M, Lasso G, Ssebide B, Nziza J, Byarugaba DK, et al. The evolutionary history of ACE2 usage within the coronavirus subgenus. Virus Evol. 2021;7:veab007.

[24] Raj VS, Smits SL, Provacia LB, van den Brand JM, Wiersma L, Ouwendijk WJ, et al. Adenosine deaminase acts as a natural antagonist for dipeptidyl peptidase 4-mediated entry of the Middle East respiratory syndrome coronavirus. J Virol. 2014;88:1834–8.

[25] Woo PC, Lau SK, Li KS, Tsang AK, Yuen KY. Genetic relatedness of the novel human group C betacoronavirus to Tylonycteris bat coronavirus HKU4 and Pipistrellus bat coronavirus HKU5. Emerg Microbes Infect. 2012;1:e35.

[26] Yang Y, Du L, Liu C, Wang L, Ma C, Tang J, et al. Receptor usage and cell entry of bat coronavirus HKU4 provide insight into bat-to-human transmission of MERS coronavirus. Proc Natl Acad Sci U S A. 2014;111:12516–21.

[27] Woo PC, Wang M, Lau SK, Xu H, Poon RW, Guo R, et al. Comparative analysis of twelve genomes of three novel group 2c and group 2d coronaviruses reveals unique group and subgroup features. J Virol. 2007;81:1574–85.

[28] Corman VM, Ithete NL, Richards LR, Schoeman MC, Preiser W, Drosten C, et al. Rooting the phylogenetic tree of middle East respiratory syndrome coronavirus by characterization of a conspecific virus from an African bat. J Virol. 2014;88:11297–303.

[29] Xiong Q, Cao L, Ma C, Tortorici MA, Liu C, Si J, et al. Close relatives of MERS-CoV in bats use ACE2 as their functional receptors. Nature. 2022;612:748–57.

[30] Huang C, Liu WJ, Xu W, Jin T, Zhao Y, Song J, et al. A Bat-Derived Putative Cross-Family Recombinant Coronavirus with a Reovirus Gene. PLoS Pathog. 2016;12:e1005883.

[31] Wu Z, Yang L, Ren X, Zhang J, Yang F, Zhang S, et al. ORF8-Related Genetic Evidence for Chinese Horseshoe Bats as the Source of Human Severe Acute Respiratory Syndrome Coronavirus. J Infect Dis. 2016;213:579–83.

[32] Schultze B, Herrler G. Recognition of cellular receptors by bovine coronavirus. Arch Virol Suppl. 1994;9:451–9.

[33] Hulswit RJG, Lang Y, Bakkers MJG, Li W, Li Z, Schouten A, et al. Human coronaviruses OC43 and HKU1 bind to 9-. Proc Natl Acad Sci U S A. 2019;116:2681–90.

[34] Yuan Y, Zu S, Zhang Y, Zhao F, Jin X, Hu H. Porcine Deltacoronavirus Utilizes Sialic Acid as an Attachment Receptor and Trypsin Can Influence the Binding Activity. Viruses. 2021;13.

[35] Wang L, Maddox C, Terio K, Lanka S, Fredrickson R, Novick B, et al. Detection and Characterization of New Coronavirus in Bottlenose Dolphin, United States, 2019. Emerg Infect Dis. 2020;26:1610–2.

